# COVID pandemic impact on hypertension management in North-East London: an observational cohort study using electronic health records

**DOI:** 10.1101/2023.08.02.23293505

**Authors:** Stuart CG Rison, Oliver Redfern, Rohini Mathur, Isabel Dostal, Chris Carvalho, Zahra Raisi-Estabragh, John P Robson

## Abstract

**Background:** The COVID19 pandemic had a major impact on primary care management of long-term conditions such as hypertension. This observational cohort study of adults with hypertension registered in 193 primary care practices in North-East London between January 2019 and October 2022 investigated the impact of the COVID19 pandemic on the treatment and control of blood pressure including demographic and social inequities.

**Method and findings:** In 224,329 adults with hypertension, the proportion with a blood pressure (BP) recorded within the preceding 1 year fell from a 91% pre-pandemic peak to 62% at the end of the pandemic lock-down phase and improved to 77% by the end of the study. The proportion with controlled hypertension (<80 years old, BP ≤140/90mmHg; 80 or more years old: ≤150/90mmHg) for the same time points was 81%, 50% and 60% respectively. Using ‘blood pressure control’ (which considered only patients with a valid blood pressure recording) as the indicator attenuated the reduction to 83%, 80% and 78% respectively.

The study used multivariable logistic analysis at four representative time points (Pre-pandemic: April 2019; Pre lockdown: April 2020; Lockdown: April 2021; Post-lockdown: April 2022) to identify temporal, clinical and demographic influences on blood pressure monitoring and control.

Pre-pandemic inequities in the management of hypertension were not significantly altered by the pandemic. Throughout the pandemic phases, in comparison to the White ethnic group, the Black ethnic group was less likely to achieve blood pressure control (ORs 0.81 [95% CI = 0.78 to 0.85, p-value<0.001] to 0.87 [95% CI = 0.84 to 0.91, p-value<0.001]). Conversely, the Asian ethnic group was more likely to have controlled blood pressure (ORs 1.09 [95% CI = 1.05 to 1.14, p-value<0.001] to 1.28 [95% CI = 1.23 to 1.32, p-value<0.001]). Younger, male, more affluent individuals, individuals with unknown or unrecorded ethnicity or those untreated were less likely to have blood pressure controlled to target throughout the study.

**Conclusion:** The COVID pandemic had a greater impact on blood pressure recording than on blood pressure control. Although recording and control have improved, these had not returned to pre-pandemic levels by the end of the study period. Ethnic inequalities in blood pressure control persisted during the pandemic and remain outstanding.

## INTRODUCTION

The COVID19 pandemic had a profound impact on the provision of primary care services and the management of long-term conditions (LTCs). Service provision in the UK and internationally, transitioned from largely face-to-face primary care appointment models to remote-consultation models accommodating reduced of direct access opportunities.[1–3] This reduced direct access was observed at multiple levels including monitoring of LTCs,[4,5] medication prescribing,[6,7] and new diagnoses of LTCs.[8–10]

The SARS-COV-19 virus also disproportionately affected certain ethnic groups both in terms of risk of infection and rate of mortality [11,12]. COVID-19 was associated with excess mortality in those of older age and with higher levels of deprivation, and highest in Black and South Asian ethnic groups; this was most marked in those with multimorbidity including cardiovascular disease, severe obesity and impaired renal function.[12]

Primary care management of pre-existing LTCs such as hypertension also worsened and may have disproportionately affected certain ethnic or other social groups.[7,13,14] Our study explores this in an ethnically diverse open cohort of individuals with hypertension monitored over 46 months spanning both the pre-pandemic, pandemic lockdown and pandemic recovery phases of the COVID19 outbreak. The objectives were: to assess the impact of the COVID pandemic on management of blood pressure of adult patients with hypertension in North-East London, and to identify any health inequities in the impact of COVID on the cohort with respect to reported ethnicity, sex, age, socio-economic status (Index of Multiple Deprivation quintile), and treatment intensity.

## METHODS

### Study cohort

The study was carried out in five contiguous North-East London localities, all former Clinical Commissioning Groups (CCGs): City and Hackney (CH), Newham (NH), Redbridge (RB), Tower Hamlets (TH) and Waltham Forest (WF) including all general practices using the EMIS electronic health record (EHR) system (EMIS health, Leeds, UK). The numbers of participating practices varied over the study period as practices opened, merged or closed.

Deidentified individual level data were collected for the first of each month from January 2019 to October 2022. Each month, the cohort comprised currently registered adults aged 18 years and older with an extant diagnosis of hypertension on the first of the month (which was considered to be the index date for that month’s cohort data). The national NHS Quality and Outcomes Framework (QOF) codeset identified hypertension excluding “hypertension resolved” (Supplementary Table S1 and https://clinicalcodes.rss.mhs.man.ac.uk/medcodes/article/203/).[15–17]

### Demographic Variables

For each individual, the following demographic data were extracted (Supplementary Table S2): Age in years on index date; Sex; home Lower Layer Super Output Areas (LSOA); Ethnic group code and study CCG. Age was banded to 18 -29 years and then to 10-year age bands to a final band of 90 years and above. Index of Multiple Deprivation was based on the 2019 Census and Lower Super Output Areas (LSOA) and used the national deprivation quintiles from quintile 1 (most deprived) to quintile 5 (least deprived).[18]

Ethnic groups were categorised according to Office of National Statistics 2001 census categories and comprised White (including White British, Irish, or White other); Black (including Black British, Caribbean, African, and other Black background); Asian (including British Asian, Bangladeshi, Pakistani, Indian and any other Asian background); Chinese and other ethnic groups (classified as Other ethnic groups); and Mixed ethnicity. [19] The Unknown ethnicity group comprised individuals with no ethnicity code recorded, individuals with unclassifiable codes and individuals with a “not stated” code.

The most recent systolic blood pressure (SBP) and diastolic blood pressure (DBP) values (in mmHg) and their entry dates were extracted. Blood pressure (BP) recordings were excluded if dating from more than 1 year prior to the index date. Recordings were also excluded if incomplete, unreliable or unfeasible blood pressures, i.e. SBP but no DBP recorded (or vice-versa), separately recorded blood pressures elements (SBP date different from DBP date), and SBP <70mmHg or SBP >=270mmHg or DBP <40mmHg or DBP >=150mmHg.

BP control was defined by NHS QOF indicators; HYP003 individuals under 80 years of age: systolic blood pressure less than or equal to 140mmHg and diastolic blood pressure less than or equal to 90mmHg); HYP007 individuals 80 years of age and older: systolic blood pressure less than or equal to 150mmHg and diastolic blood pressure less than or equal to 90mmHg.[20]

Medicines prescribed in the 6 months up to and including the index date were considered for eight classes of antihypertensive medication (Supplementary Table S2): i) ACE inhibitors/Angiotensin Receptor Blockers, ii) Beta-blockers, iii) Potassium-sparing diuretics, iv) Calcium channel blockers, v) Thiazide-type and thiazide-like diuretics, vi) Centrally-acting antihypertensives, vii) Alpha-blockers and viii) Loop diuretics. The number of different classes prescribed were grouped into categoric treatment intensities, i.e. individuals on 0, 1, or 2 or more antihypertensive medication classes.[16,17]

### Outcomes

Three binary outcome variables of blood pressure management were considered.[16,17]

1. BLOOD_PRESSURE_RECORDED: a valid blood pressure recorded within 12 months of the index date.
2. HYPERTENSION_CONTROLLED: a blood pressure within the QOF age-adjusted target. All individuals without a blood pressure within 12 months of the index date were considered to be above target blood pressure and not controlled.
3. BLOOD_PRESSURE_CONTROLLED: Individuals with blood pressure recorded within 12 months of the index date who had blood pressure within the QOF age-adjusted target. This measure excluded people without a record of blood pressure within 12 months.

### Study Phases

The 1^st^ April 2019, 2020, 2021 and 2022 were considered to be the representative index date for the following pandemic-related phases: pre-pandemic, pandemic pre-lockdown, pandemic lockdown and pandemic recovery respectively.[21] Herein, these phases are described by name or by the year of the phase.

### Statistical methods

Data were processed, aggregated and validated, descriptive statistics derived, and outcome indicator trajectories graphed (as percentage of cohort individuals meeting the indicator criteria). Correlations between indicator trajectories were measured using Pearson correlation coefficients.

Unadjusted and adjusted analyses (for ethnic group, sex, age, IMD quintile and treatment intensity) multivariable logistic regression were completed for each outcome indicator. Interaction terms between pandemic phases (as categorical year variables as described above) and other variables were included in both models to calculate odds ratios (ORs) per pandemic phase.

All analyses were performed using Python (version 3.9.1) and R (version 4.0.5). Forest plots were generated using the forestplot Python package (version 0.2.0).[22]

## RESULTS

### Study cohort and demographic variables

There were 199 GP practices in the five North-East London study localities (CCGs) and 193 of these were included in the study with a total adult population of approximately 1.4 million individuals. Practice numbers varied from 187 to 193 depending on the study month as a result of practice openings, closures or mergers.

Over the 46 months of the study, data from 224,329 individuals were considered, with an average of 33.5 patient months of follow-up (Figure 1 and Table 1). 113,255/224,329 [50.5%] of individuals were present throughout the 46 months of the study. 215,219/224,329 [95.9%] of individuals had at least one valid blood pressure recording throughout the study period. (Supplementary Table S3).

**Figure 1:**
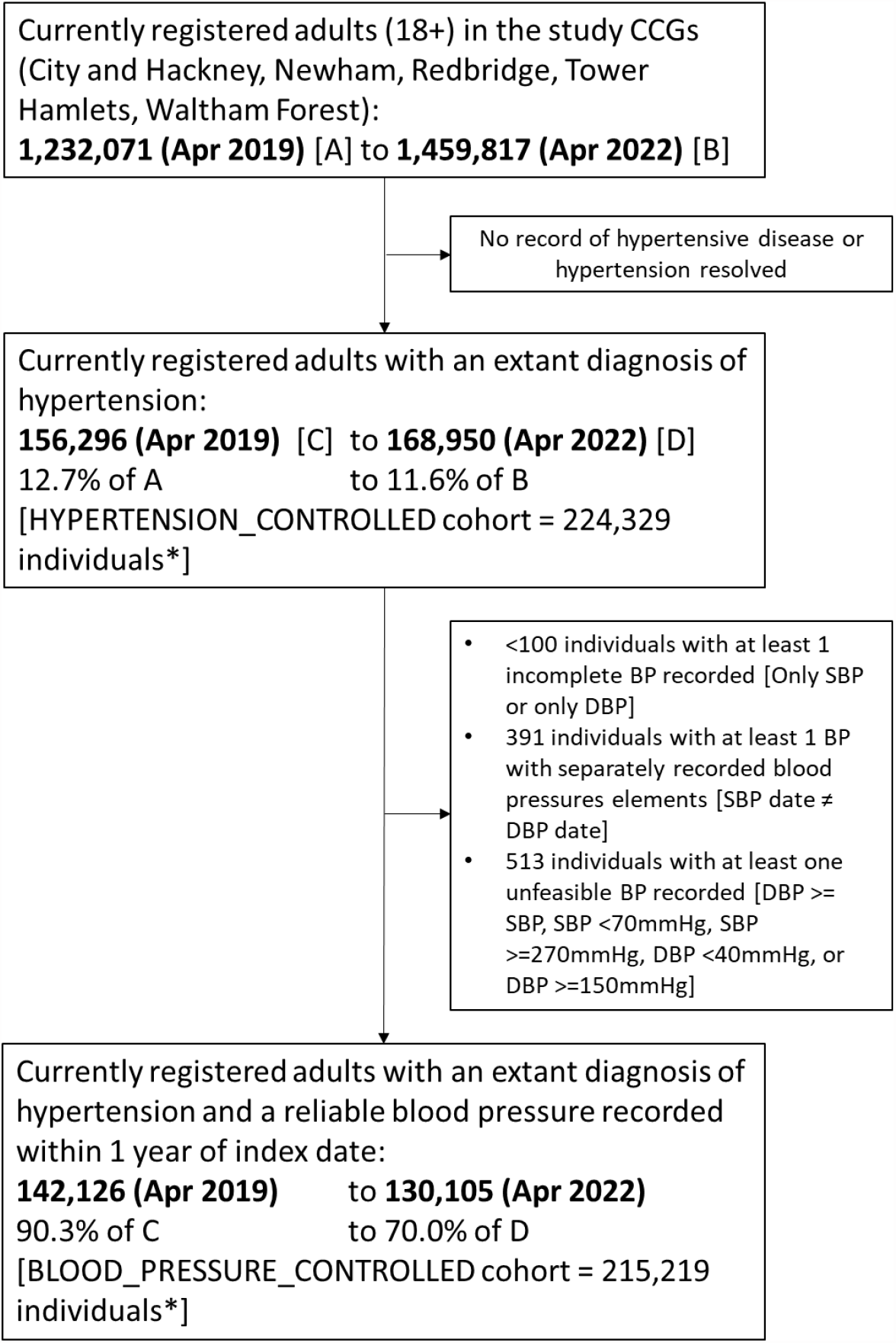
Study open cohort flowchart. The study ran from 01/01/2019 to 01/10/2022 with April 2019 and April 2022 representing the pre-pandemic and post-lockdown cohort respectively. SBP = Systolic Blood Pressure, DBP = Diastolic Blood Pressure. *Unique cohort members over the whole study period.

**Table 1:** Characteristics of patients included in the analysis. Characteristics are derived from the first presentation of an individual in the open cohort except for time in study which considers individuals throughout the study period (n= 224,329 adults with a diagnosis of hypertension). For categorical characteristics, the most common category is shown in bold. BP = Blood Pressure, SD = Standard Deviation.

|  |  |  |
| --- | --- | --- |
| Individual patients [n] | 224,329 |  |
| Age (years): mean [SD] | 61.3 [14.6] |  |
| Systolic BP (mmHg): mean [SD] | 133.6 [14.3] |  |
| Diastolic BP (mmHg): mean [SD] | 79.0 [10.2] |  |
| Time in study (months): mean [SD; min.; max.; mode] | 33.5 [15.7; 1; 46; 46] |  |
| Sex: n [%] |  |  |
| Female | 113,604 [50.6%] |  |
| Male | 110,725 [49.4%] |  |
| Ethnicity: n [%] |  |  |
| White | 82,624 [36.8%] |  |
| Asian or Asian British | 68,701 [30.6%] |  |
| Black or Black British | 46,111 [20.6%] |  |
| Unknown | 12,883 [5.7%] |  |
| No ethnicity code recorded | 7,872 [3.5%] |  |
| Unclassified code | 3,504 [1.6%] |  |
| “Not stated” code | 1,507 [0.7%] |  |
| Other Ethnic Group | 9,255 [4.1%] |  |
| Mixed | 4,755 [2.1%] |  |
| Age distribution: n [%] |  |  |
| (18-30] | 2,578 [1.1%] |  |
| (30-40] | 12,764 [5.7%] |  |
| (40-50] | 33,333 [14.9%] |  |
| (50-60] | 55,856 [24.9%] |  |
| (60-70] | 53,850 [24.0%] |  |
| (70-80] | 38,119 [17.0%] |  |
| (80-90] | 22,802 [10.2%] |  |
| (90-120] | 5,027 [2.2%] |  |
| Index of Multiple Deprivation (IMD) Quintile: n [%] |  |  |
| Q1 (most deprived) | 59,368 [26.5%] |  |
| Q2 | 104,568 [46.6%] |  |
| Q3 | 36,766 [16.4%] |  |
| Q4 | 16,857 [7.5%] |  |
| Q5 (least deprived) | 6,634 [3.0%] |  |
| Q0 (Unrecorded) | 136 [0.1%] |  |
| Number of antihypertensive medications: n [%] |  |  |
| 0 | 39,591 [17.6%] |  |
| 1 | 82,642 [36.8%] |  |
| 2 | 61,305 [27.3%] | 2+:<br>102,096<br>[45.5%] |
| 3 | 28,354 [12.6%] |  |
| 4 | 9,659 [4.3%] |  |
| 5+ | 2,778 [1.2%] |  |

Characteristics of the cohort are described in Table 1. 96.5% of individuals had a self-reported ethnicity code recorded and an ONS ethnicity group was assigned to 94.3% of individuals (accounting for individuals with no ethnicity code, “not stated” ethnicity codes and ethnicity codes which could not be mapped to an ONS ethnicity group).

The cohort make-up in terms of ethnicity groups, ages, sex, IMD quintiles and treatment intensities remained largely unchanged over the study period (supplementary Figure S1).

### Outcomes

The outcome variables were plotted as percentages over the study period (Figure 2). The trendline for blood pressure recordings (BLOOD_PRESSURE_RECORDED) showed a large fall in the pandemic lockdown phase from 89% in April 2020 to 62% at the end of the lockdown phase, paralleled by a fall in HYPERTENSION_CONTROLLED from 73% to 50%. Both indicators then showed a marked rise in the pandemic recovery phase but not to pre-pandemic levels. The BLOOD_PRESSURE_RECORDED and HYPERTENSION_CONTROLLED indicators were highly correlated (Pearson correlation coefficient (r)= 0.99; p-value <0.001). In contrast, the BLOOD_PRESSURE_CONTROLLED indicator exhibited a smaller fall during lockdown, from a peak of 83% in April 2020 to 80% at the end of the study. Furthermore, there was no significant correlation between BLOOD_PRESSURE_CONTROLLED and BLOOD_PRESSURE_RECORDED (r= 0.20; p-value 0.18).

**Figure 2:**
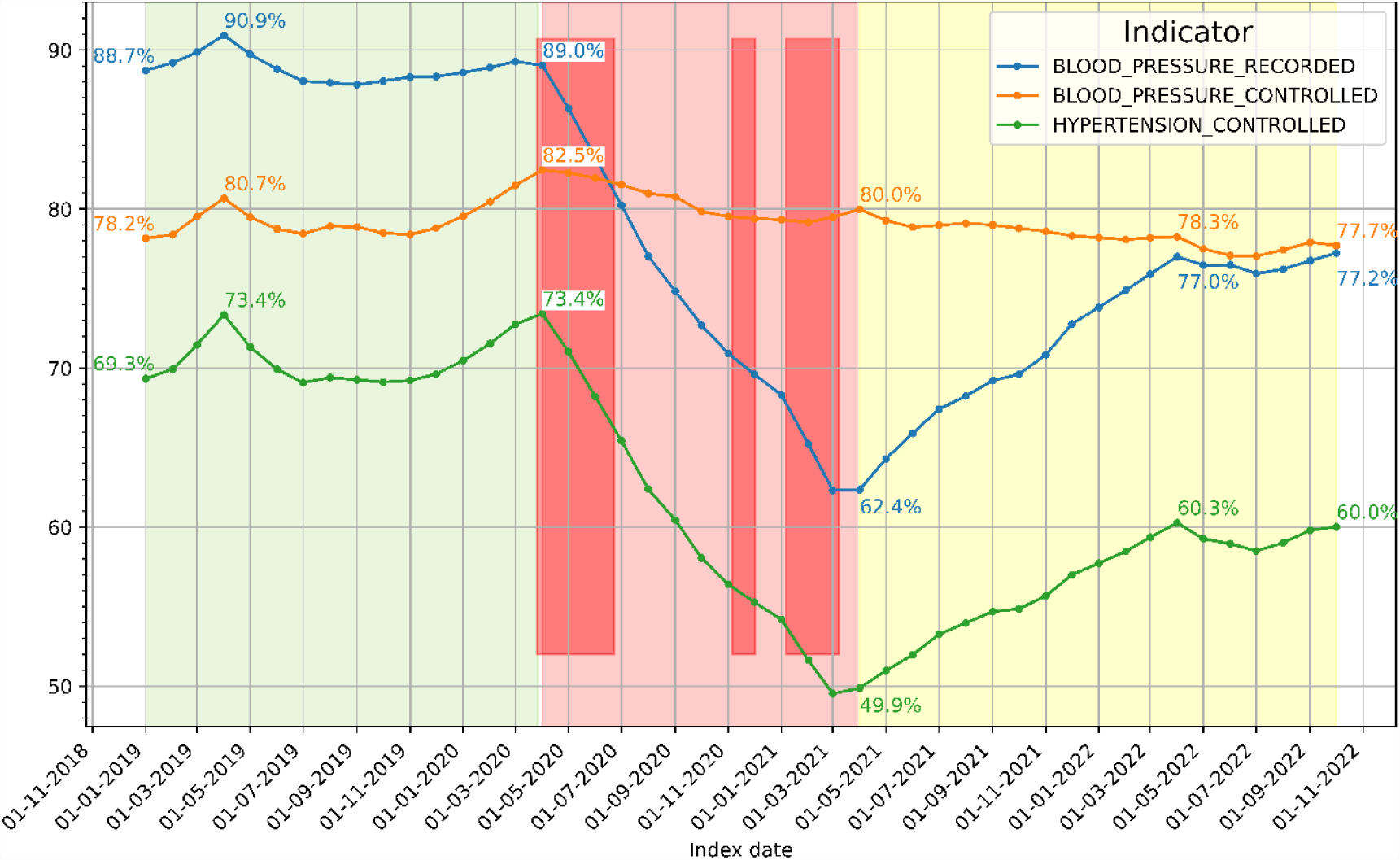
Changes in the BLOOD_PRESSURE_RECORDED, BLOOD_PRESSURE_CONTROLLED, and HYPERTENSION_CONTROLLED indicators over the course of the study. The graph is divided in three phases: pre-pandemic (green area), pandemic-lockdown (light red area) and pandemic-recovery (yellow area). The three darker red rectangles represent the three England-wide COVID lockdowns. Percentages are printed for the April 2019, 2020, 2021 and 2022 outcomes as well as for the first and last months of the study.

One study locality (the City and Hackney CCG) launched a blood pressure recording initiative in August 2020. To assess the impact of the intervention, the indicator trend plots were subdivided by study locality as shown in Supplementary Figure S2. In the intervention locality, the Pearson coefficients between BLOOD_PRESSURE_RECORDED and the HYPERTENSION_CONTROLLED was 0.96 (p-value<0.001). The correlation between BLOOD_PRESSURE_RECORDED and BLOOD_PRESSURE_CONTROLLED was not significant (r= 0.62; p-value=3.90). The initiative did increase the percentage of contemporary blood pressure recording in City and Hackney (BLOOD_PRESSURE_RECORDED) with a parallel improvement in the HYPERTENSION_CONTROLLED indicator, but this did not translate into a higher percentage of patient with blood pressures controlled to target (i.e. there was no associated rise in the BLOOD_PRESSURE_CONTROLLED outcomes in City and Hackney).

### Study phases

#### Pre-pandemic phase analyses

Analysis of the pre-pandemic cohort has been accepted for publication having been pre-submitted to medRxiv.[16,17] In comparison to the White ethnicity group, the Black ethnicity group was less likely to have BLOOD_PRESSURE_CONTROLLED (OR 0.87, 95% CI = 0.84-0.91). The Asian ethnicity group was more likely to have BLOOD_PRESSURE_CONTROLLED (OR 1.28, 95% CI = 1.23-1.32). Ethnicity group differences in hypertension control could not be explained by the likelihood of having a recent blood pressure recording, nor by treatment intensity differences. Older adults and individuals living in more deprived areas were more likely to have controlled hypertension than younger patients and individuals in more affluent areas.

#### Variation between study period phases – ethnic variations

Radar charts for outcomes by ethnic group are shown in Figure 3. Forest plots including 95% confidence intervals and p-values are shown in supplementary Figure S3; the whole model variables are shown in supplementary Figure S4.

**Figure 3:**
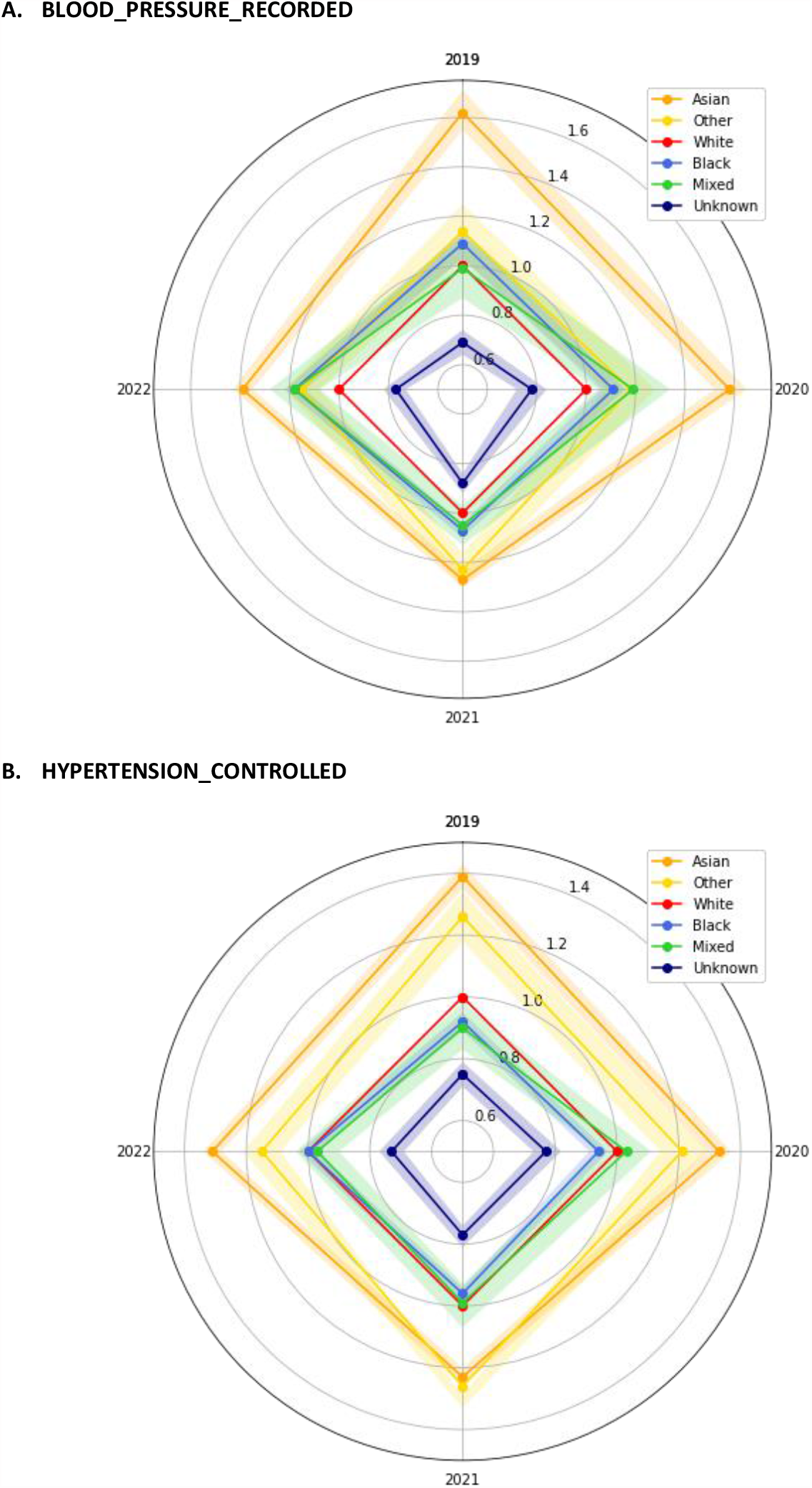
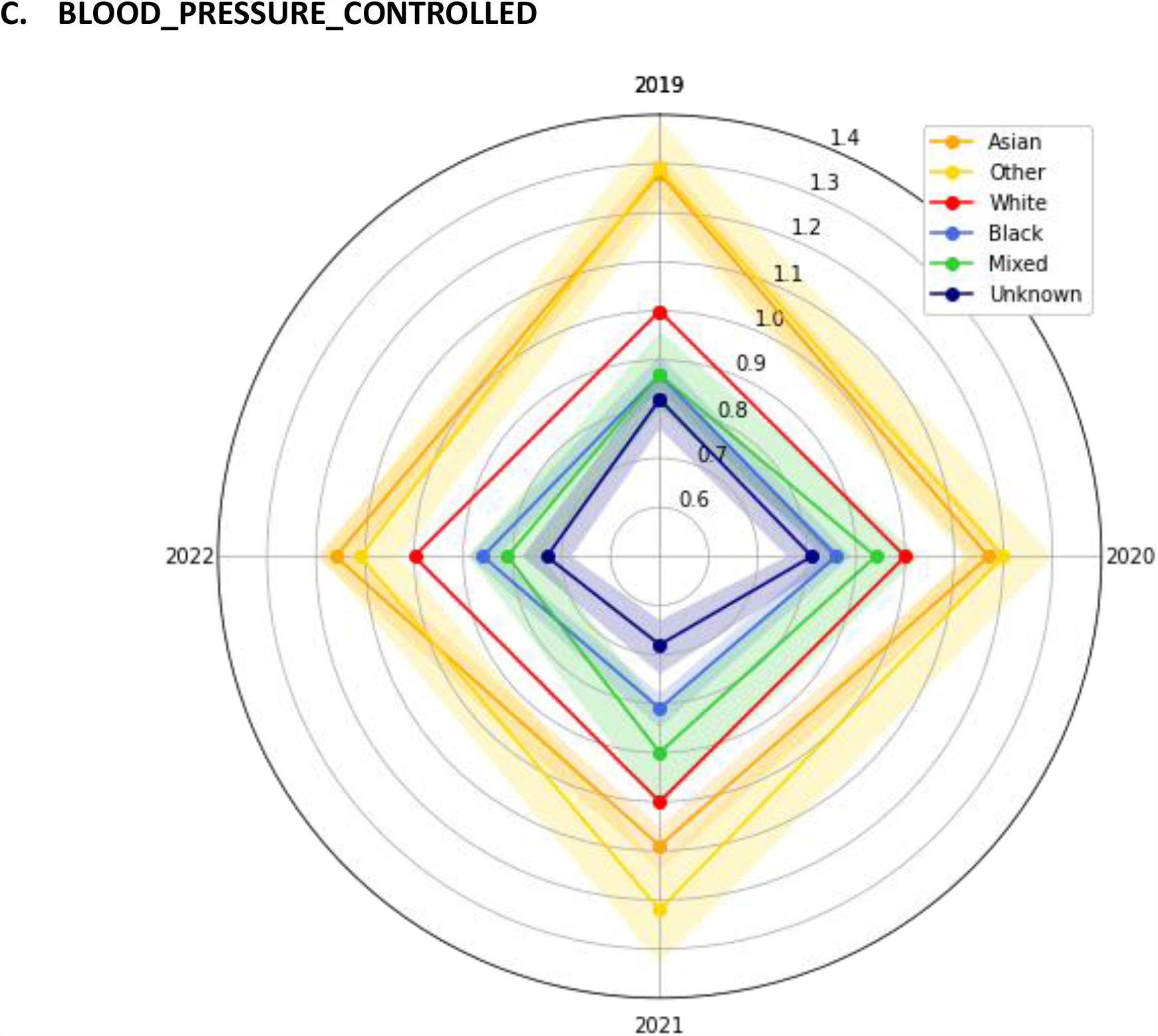
Radar charts of odd ratios (clockwise from 12 o’clock: April 2019, April 2020, April 2021, April 2022) by study ethnicity group: A. BLOOD_PRESSURE_RECORDED, B. HYPERTENSION_CONTOLLED, C. BLOOD_PRESSURE_CONTROLLED. 95% confidence intervals are shown (shaded areas). The White ethnicity group (red line) is the reference group and therefore plotted with an OR of 1.0 and no confidence interval.

The radar charts confirm that, relative to the White ethnic group and regardless of the pandemic phase, the Black ethnic group always had worse HYPERTENSION_CONTROLLED and BLOOD_PRESSURE_CONTROLLED (i.e. the Black ethnic group polygon (light blue) is fully circumscribed by the White ethnic group polygon (red)) (Figure 3). Conversely, the Asian ethnic group always had larger odd ratios than the White ethnic group.

For the BLOOD_PRESSURE_RECORDED indicator, the Asian ethnic group polygon fully circumscribes all the other ethnic groups. However, the White ethnic group was less likely than the Black ethnic group to have a recently recorded blood pressure throughout the study phases. Regardless of the indicator considered, the relatively small and heterogenous Unknown ethnicity group (navy blue inner polygon) always had the worst outcomes (i.e. smallest odds ratios (ORs)).

**<u>Figure</u>** and the forest plots of the ORs for the three indicators by ethnicity group (supplementary Figure S3) clarify the relative impact of the pandemic phases on these groups. Using the White ethnic group as the comparator and April 2019 ORs as the baseline, subsequent OR increases or decreases indicate improving or worsening performance respectively. For example, for BLOOD_PRESSURE_RECORDED in the Asian ethnic group, from a baseline OR of 1.28, the group performed worse in 2020 (OR 1.17), worse still in 2021 (OR 1.09) and partially recovered in 2022 (OR 1.16). The Black ethnic group had the same pattern of worsening and improvement. During the pandemic (2021), both the Asian and Black ethnic group ORs fell further than in the White ethnic group, but these changes did not affect the overall picture: throughout the study, the Asian ethnic group always performed better than the White ethnic group and the Black ethnic group always worse in both indicators of blood pressure control.

#### Variation between study period phases – other variations

Male, younger, and more affluent individuals were less likely to have a recent blood pressure recorded or controlled hypertension.[16] These differences were also present throughout the study (Supplementary Figure S4).

Treatment intensity was relevant to the BLOOD_PRESSURE_RECORDED indicator. In the pre-pandemic phase, compared to individuals on no antihypertensives, individuals on 1, or 2 or more antihypertensives were markedly more likely to have a blood pressure recorded, ORs: 7.8 [7.5-9.2; p-value <0.001] and 10.0 [9.6-10.5; p-value <0.001] respectively. This effect was less marked in later phases with ORs ranging from 3.3 to 8.6 (Supplementary Table S4). The effect of treatment intensity was also seen with the other indicators, but it was less prominent. For both HYPERTENSION_CONTROLLED and BLOOD_PRESSURE_CONTROLLED, untreated individuals were, predictably, less likely to have controlled blood pressure than individuals on one or more antihypertensives.

## DISCUSSION

### Summary

This study followed a large open-cohort of 224,329 adults with hypertension from 193 GP practices in North-East London over 46 months including the pre-pandemic, the pandemic pre-lockdown, the pandemic lockdown and the pandemic recovery phases.

The national QOF performance indicators for hypertension control yielded unduly low estimates of control of blood pressure because they conflate lack of recording with lack of control –a situation well illustrated by the City and Hackney lockdown blood pressure recording initiative which effected a notable improvement in the percentage of patient with a blood pressure recorded within a year, and consequently in the QOF HYP003 and HYP007 indicators, but made little change to percentage of controlled blood pressures *per se*. Thus, the pandemic impacted all our indicators of hypertension management, but the impact on recording of blood pressure was much more marked than reductions in the control of blood pressure. Furthermore, more than one year after the pandemic lockdowns, levels of control of blood pressure have improved but have yet to be restored to pre-pandemic levels and ethnic inequalities are persistent.

Existing ethnic variations persisted in all pandemic phases. Both before, during and “after” the pandemic, the Black ethnic group was less likely to have controlled hypertension, and the Asian ethnic group more likely to have controlled hypertension, than the White ethnicity group. This was not due to less frequent recording of blood pressure in the Black ethnic group, nor was it due to an association with age or deprivation. Indeed, patients in the most deprived IMD quintile were more likely to have controlled hypertension than patients in less deprived IMD quintiles. Furthermore, this inequity was present regardless of the treatment intensity and the Black ethnic group was as intensively treated as other ethnicity groups. Control of hypertension was also less likely in male and in younger individuals throughout the study period.

Individuals in the Unknown ethnicity group (i.e. ethnicity not recorded, recorded as “not stated” or with an unclassifiable ethnicity code) and individuals not on any antihypertensives fared worst. In both cases, it is likely that these individuals had fewer contacts with healthcare professionals and may include some individuals who had in fact left their practice. More broadly, ethnicity recording – or more saliently the lack thereof– is a marker of healthcare disengagement.

At the start of the study (January 2019), BLOOD_PRESSURE_CONTROLLED was 78.2%, by the end of the study (October 2022), it was 77.7%. Therefore, although the pandemic had a profound impact on the recording of blood pressures, it appears to have had limited impact on this indicator of hypertension management. However, this before and after comparison conceals a more recent and sizeable deterioration as the pre-lockdown peak was 82.5% in April 2020.

The relationship of treatment intensity to BLOOD_PRESSURE_RECORDED was particularly marked and was 10 times higher for individuals on 2 or more medications than for untreated individuals. Untreated individuals were more likely to be younger, male or be of unknown or unrecorded ethnicity representing patients who may find access to primary care difficult, or be less willing to engage with health care providers. Untreated individuals represent an under-monitored group.

The COVID pandemic effected a radical change in the provision of primary care service and reduced opportunities for recording of risk factors and optimising management. In North-East London, significant efforts are being made to restore performance of health provision but our data show that despite substantial improvements, more than a year after the pandemic, recovery to pre-pandemic levels has yet to be achieved. Restoration of blood pressure recording is a key element in optimising disease management, and targeted improvements in management of hypertension in the Black ethnic group remains an outstanding priority.[23]

### Strengths and limitations

This large study offers a unique perspective on the impact of the COVID pandemic in the management of a major long-term condition. The study cohort was unselected, including 193 (97%) of the 199 practices in the 5 localities (CCGs) considered in the study and with 94.3% recording of self-reported ethnicity in an ethnically diverse population with a broad representation of the principal ethnicity groups discussed.

Treatment changes within 6 months would not be captured accurately, possibly overestimating treatment intensity in a small number of patients. Area level measures of deprivation such as IMD quintile reduce gradients between extremes of deprivation. In addition, during the pandemic, the most affluent may have been away from their London residences for prolonged periods or accessing alternative health services contributing to the apparent poor management in this group.

Although our study considered multiple demographic factors, the inequities identified in our study might be due to other confounding factors such as adherence to treatment, treatment escalation, or ethnic variability in the prevalence of resistant hypertension.[24–29] The study may also have been affected by the impact of the pandemic on hypertension case finding; a UK wide study found that nearly 500,000 fewer people than expected started taking blood pressure lowering medication between March 2020 and July 2021.[10]

### Comparison with existing literature

Comparing the 2020 and 2021 CVDPREVENT audits, the authors reported similar findings. The pandemic caused a similar reduction across sex, age and ethnicity group in recording of blood pressures (CVDP004HYP).[30] There was little change in differentials by age, deprivation and ethnicity group control of blood pressure in those with recent blood pressure recorded and the authors concluded that the impact was largely on recording of blood pressure rather than control.[30]

Findings internationally were similar. A large study from 24 US health systems from 2017 to 2020 (PCORnet study) investigated indicators of hypertension control including the percentage of patients with a blood pressure <140/90mmHg.[31] Findings for this indicator paralleled those in our study: patients of Asian ethnicity always fared better, and patients of Black ethnicity always fared worse than patients of White ethnicity, despite higher use of follow-up visits in the Black ethnicity group.

Weighted average BP control was uncontrolled for 60.5% of individuals in the PCORnet 2019 pre-pandemic cohort and dropped over 7.2 percentage points for the 2020 pandemic cohort. In our study, the equivalent BOOD_PRESSURE_CONTROL indicator peaked at 82.5% at the end of the pre-pandemic phase, fell to 80% by the end of the lockdown phase (a 2.5 percentage point fall) and, by the end of the study was at 77.7% (a 4.8% percentage fall).

### Implications for research and practice

The study highlighted the importance of metric choice in understanding control of hypertension in a population. QOF indicators such as HYP003 and HYP007 may serve as a suitable “performance” indicator but a better reflection of the control of hypertension requires consideration of both recording of blood pressure and control in those recorded.

Certain groups of individuals consistently fare worse and may warrant targeted intervention including the Black ethnic group, younger and untreated patients. Our study also suggests individuals diagnosed with hypertension but not on any treatment are less likely to have their blood pressure monitored than their treated counterparts –increasing frequency of monitoring in these individuals might be beneficial. Lastly, although people with missing ethnicity are a relatively small group, they nevertheless accounted for almost 6% of our cohort and management of hypertension was particularly poor in this group –understanding both systemic and patient-level obstacles to ethnicity recording may well also identify obstacles to effective management of long-term conditions for this community. As always, competing priorities and resource restriction complicate such targeted interventions, but health economic modelling in North East London has already demonstrated that improved blood pressure control can cost-effectively reduce cardiovascular risk and improve life expectancy. [32]

## Data Availability

All data produced in the present study are available upon reasonable request to the authors

## ETHICS APPPOVAL

This study is based on de-identified information obtained from routinely compiled general practitioner electronic health records. All participating GP practices consented to the use of their anonymised patient data for research and development for patient benefit.

Based on the NHS Health Research Authority Questionnaire (http://www.hra-decisiontools.org.uk/ethics/), research ethics approval was not required for this project as patient level data are anonymised, and only aggregated patient data are reported in this study.

This was confirmed by the Chair of the North-East London Strategic Information Governance Network.

## COMPETING INTERESTS

The authors have declared no competing interests.

## ACKNOWLEDGMENTS

We are grateful to the general practitioners and their practice teams for allowing the use of their patient records, to the Clinical Effectiveness Group for providing access to their curated high-quality dataset and to the population in North-East London from whom the data are derived. The authors wish to thank staff at CEG for supporting practices with guidance and data entry tools which support this project.

We are grateful for Lucas Shen’s help with his forestplot Python package.

## FUNDING

This work was supported by Barts Charity and Health Data Research UK, an initiative funded by UK Research and Innovation, Department of Health and Social Care (England) and the devolved administrations, and leading medical research charities. OR is supported by the National Institute for Health Research (NIHR) and a Drayson research fellowship. RM is supported by Barts Charity (MGU0504). ZR-E is supported by an NIHR Integrated Academic Training programme and her Academic Clinical Lectureship post and is also supported by a British Heart Foundation Clinical Research Training Fellowship (FS/17/81/33318). CC is supported by an NIHR School of Primary Care Research Clinical Fellowship Queen Mary University of London. The views expressed are those of the authors and not necessarily those of the NIHR or the Department of Health and Social Care.

## Notes

### Author Declarations

The clinical effectiveness group (CEG) has the written consent of all practices in the study area to use pseudonymised patient data for audit and research for patient benefit. The CEG is the data processor, and the General Practices in the study are the data controllers. The researchers adhere to the data protection principles of the Data Protection Act 2018, and all data was managed according to UK NHS information governance requirements. All data were pseudonymised and are only presented in aggregate form. The NHS Health Research Authority toolkit (http://www.hra-decisiontools.org.uk/ethics/) identified that Research Ethics Approval was not required for this project. This was confirmed by the Chair of the North East London Strategic Information Governance Network.

## REFERENCES

1 Lim J, Broughan J, Crowley D, et al. COVID-19’s impact on primary care and related mitigation strategies: A scoping review. Eur J Gen Pract 2021;27:166–75. doi:10.1080/13814788.2021.1946681

2 Patel SY, Mehrotra A, Huskamp HA, et al. Trends in Outpatient Care Delivery and Telemedicine During the COVID-19 Pandemic in the US. JAMA Intern Med 2021;181:388–91. doi:10.1001/jamainternmed.2020.5928

3 British Medical Association. BMA COVID review 3: Delivery of healthcare during the pandemic. 2022.

4 Maddock J, Parsons S, Di Gessa G, et al. Inequalities in healthcare disruptions during the COVID-19 pandemic: evidence from 12 UK population-based longitudinal studies. BMJ Open 2022;12:e064981. doi:10.1136/bmjopen-2022-064981

5 McGreevy A, Soley-Bori M, Ashworth M, et al. Ethnic inequalities in the impact of COVID-19 on primary care consultations: a time series analysis of 460,084 individuals with multimorbidity in South London. BMC Med 2023;21:26. doi:10.1186/s12916-022-02720-7

6 Bankhead CR, Lay-Flurrie S, Nicholson BD, et al. Changes in cardiovascular disease monitoring in English primary care during the COVID-19 pandemic: an observational cohort study. medRxiv 2020;:2020.12.11.20247742. doi:10.1101/2020.12.11.20247742

7 Carr MJ, Wright AK, Leelarathna L, et al. Impact of COVID-19 restrictions on diabetes health checks and prescribing for people with type 2 diabetes: a UK-wide cohort study involving 618 161 people in primary care. BMJ Qual Saf Published Online First: 12 October 2021. doi:10.1136/bmjqs-2021-013613

8 Williams R, Jenkins DA, Ashcroft DM, et al. Diagnosis of physical and mental health conditions in primary care during the COVID-19 pandemic: a retrospective cohort study. Lancet Public Heal 2020;5:e543–50. doi:10.1016/S2468-2667(20)30201-2

9 Grant MP, Helsper CW, Stellato R, et al. The Impact of the COVID Pandemic on the Incidence of Presentations with Cancer-Related Symptoms in Primary Care. Cancers (Basel) 2022;14. doi:10.3390/cancers14215353

10 Dale CE, Takhar R, Carragher R, et al. The impact of the COVID-19 pandemic on cardiovascular disease prevention and management. Nat Med 2023;29:219–25. doi:10.1038/s41591-022-02158-7

11 McQueenie R, Foster HME, Jani BD, et al. Multimorbidity, polypharmacy, and COVID-19 infection within the UK Biobank cohort. PLoS One 2020;15:e0238091. doi:10.1371/journal.pone.0238091

12 Williamson EJ, Walker AJ, Bhaskaran K, et al. Factors associated with COVID-19-related death using OpenSAFELY. Nature 2020;584:430–6. doi:10.1038/s41586-020-2521-4

13 Costello RE, Tazare J, Piehlmaier D, et al. Ethnic differences in the indirect effects of the COVID-19 pandemic on clinical monitoring and hospitalisations for non-COVID conditions in England: a population-based, observational cohort study using the OpenSAFELY platform. EClinicalMedicine 2023;61:102077. doi:10.1016/j.eclinm.2023.102077

14 Karagiannidis AG, Theodorakopoulou MP, Ferro CJ, et al. Impact of public restrictive measures on hypertension during the COVID-19 pandemic: existing evidence and long-term implications. Clin Kidney J 2023;16:619–34. doi:10.1093/ckj/sfac235

15 NHS Digital. Quality and Outcomes Framework (QOF) business rules v42 2019-2020 baseline release. 2019. https://digital.nhs.uk/data-and-information/data-collections-and-data-sets/data-collections/quality-and-outcomes-framework-qof/quality-and-outcome-framework-qof-business-rules/quality-and-outcomes-framework-qof-business-rules-v42-2019-2020-baseline-releas x(accessed 1 Jul 2020).

16 Rison SC, Redfern O, Dostal I, et al. Inequities in hypertension management: an observational cross-sectional study in North-East London using electronic health records. medRxiv Published Online First: 2023. doi:10.1101/2023.03.14.23286419

17 Rison SC, Redfern O, Dostal I, et al. Inequities in Hypertension Management: an observational cross-sectional study in North-East London using electronic health records. Br J Gen Pract 2013;In press.

18 Ministry of Housing Communities and Local Government. English indices of deprivation 2019. 2019. https://www.gov.uk/government/statistics/english-indices-of-deprivation-2019

19 HM Government. List of ethnic groups. Ethn. facts Fig. - Style Guid. 2021. https://www.ethnicity-facts-figures.service.gov.uk/style-guide/ethnic-groups (accessed 24 Jul 2020).

20 NHS Digital. Quality and Outcomes Framework (QOF) business rules v47.0 2022-2023 baseline release. 2022. https://digital.nhs.uk/data-and-information/data-collections-and-data-sets/data-collections/quality-and-outcomes-framework-qof/quality-and-outcome-framework-qof-business-rules/quality-and-outcomes-framework-qof-business-rules-v47.0-2022-2023-baseline-rele

21 Brown J, Kirk-Wade E. Coronavirus: A history of English lockdown laws. House of Commons Library Briefing Paper no. 6964. 2021. https://researchbriefings.files.parliament.uk/documents/CBP-9068/CBP-9068.pdf

22 Shen L, Shapiro A. LSYS/forestplot: v0.2.0. 2022. doi:10.5281/zenodo.7270258

23 Imperial College Health Partners. Hypertension project: OneLondon Pathfinder programme - Evaluation. 2023. https://selondonccg.nhs.uk/wp-content/uploads/2023/02/Hypertension-Pathfinder-Evaluation-report-FINAL.pdf

24 Roberie DR, Elliott WJ. What is the prevalence of resistant hypertension in the United States? Curr Opin Cardiol 2012;27:386–91. doi:10.1097/HCO.0b013e328353ad6e

25 Smith SM, Gurka MJ, Winterstein AG, et al. Incidence, prevalence, and predictors of treatment-resistant hypertension with intensive blood pressure lowering. J Clin Hypertens (Greenwich) 2019;21:825–34. doi:10.1111/jch.13550

26 Eastwood S V., Hughes AD, Tomlinson L, et al. Ethnic differences in hypertension management, medication use and blood pressure control in UK primary care, 2006–2019: A retrospective cohort study. Lancet Reg Heal - Eur 2022;0:100557. doi:10.1016/j.lanepe.2022.100557

27 Lackland DT. Racial differences in hypertension: implications for high blood pressure management. Am J Med Sci 2014;348:135–8. doi:10.1097/MAJ.0000000000000308

28 Redmond N, Baer HJ, Hicks LS. Health behaviors and racial disparity in blood pressure control in the national health and nutrition examination survey. Hypertens (Dallas, Tex 1979) 2011;57:383–9. doi:10.1161/HYPERTENSIONAHA.110.161950

29 Aggarwal R, Chiu N, Wadhera RK, et al. Racial/Ethnic Disparities in Hypertension Prevalence, Awareness, Treatment, and Control in the United States, 2013 to 2018. Hypertens (Dallas, Tex 1979) 2021;78:1719–26. doi:10.1161/HYPERTENSIONAHA.121.17570

30 Office for Health Improvement & Disparities (OHID) and NHS Benchmarking Network. CVDPREVENT Second Annual Audit Report. 2022.

31 Chamberlain AM, Cooper-DeHoff RM, Fontil V, et al. Disruption in Blood Pressure Control With the COVID-19 Pandemic: The PCORnet Blood Pressure Control Laboratory. Mayo Clin Proc Published Online First: January 2023. doi:10.1016/j.mayocp.2022.12.024

32 Wu R, Rison SCG, Raisi-Estabragh Z, et al. Gaps in antihypertensive and statin treatments and benefits of optimisation: A modelling study in a 1 million ethnically diverse urban population in UK. BMJ Open 2021;11. doi:10.1136/bmjopen-2021-052884

