## Supplementary material for "COVID pandemic impact on hypertension management in North-East London: an observational cohort study using electronic health records": Figure S1

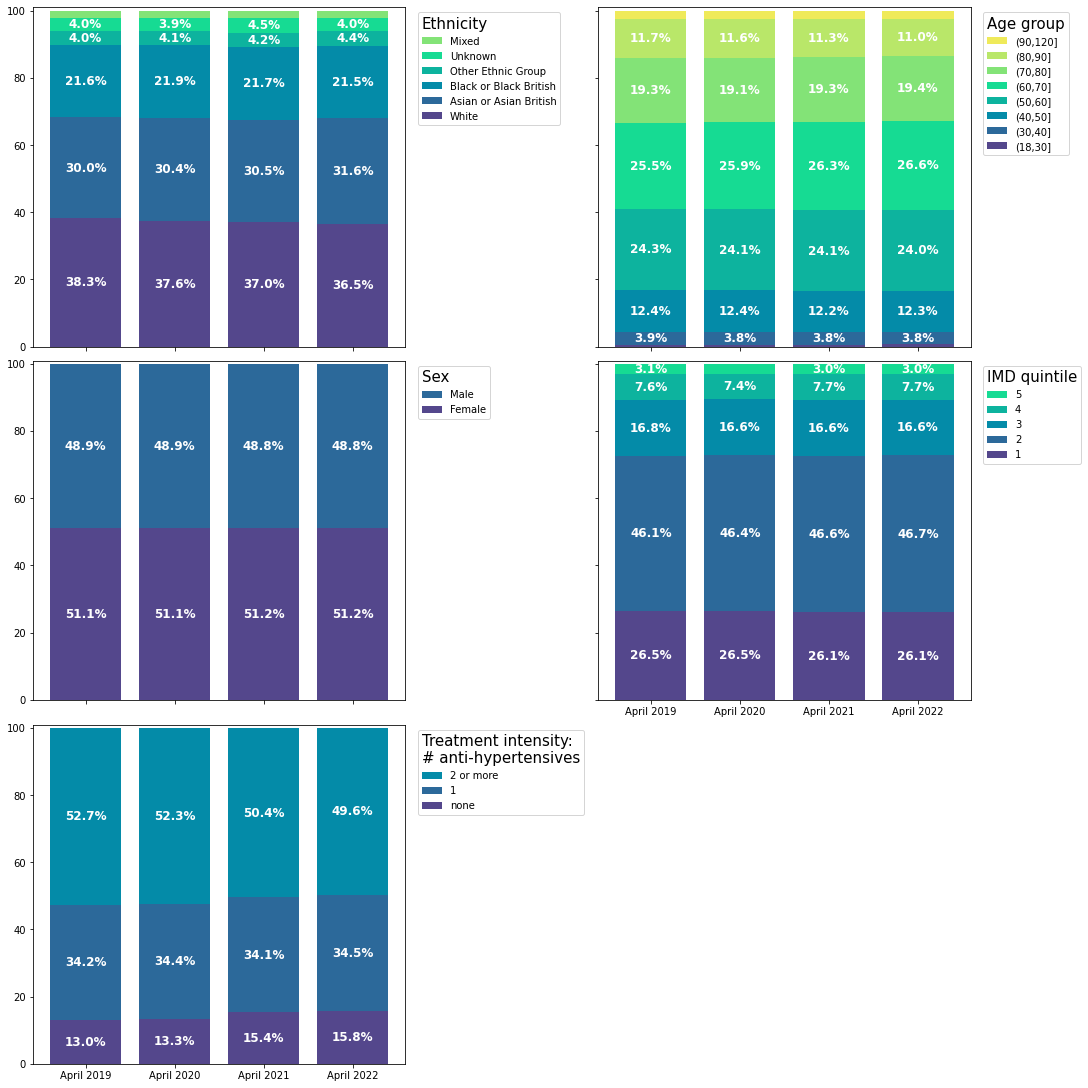

**Figure S1:** Cohort characteristic (ethnicity, age, sex, IMD quintile and treatment intensity) as a percentage of the cohort for the cohort on 1st April 2019,2020,2021 and 2022). Percentages under 3.0% are not labelled.
