## Supplementary material for "COVID pandemic impact on hypertension management in North-East London: an observational cohort study using electronic health records": Figure S2

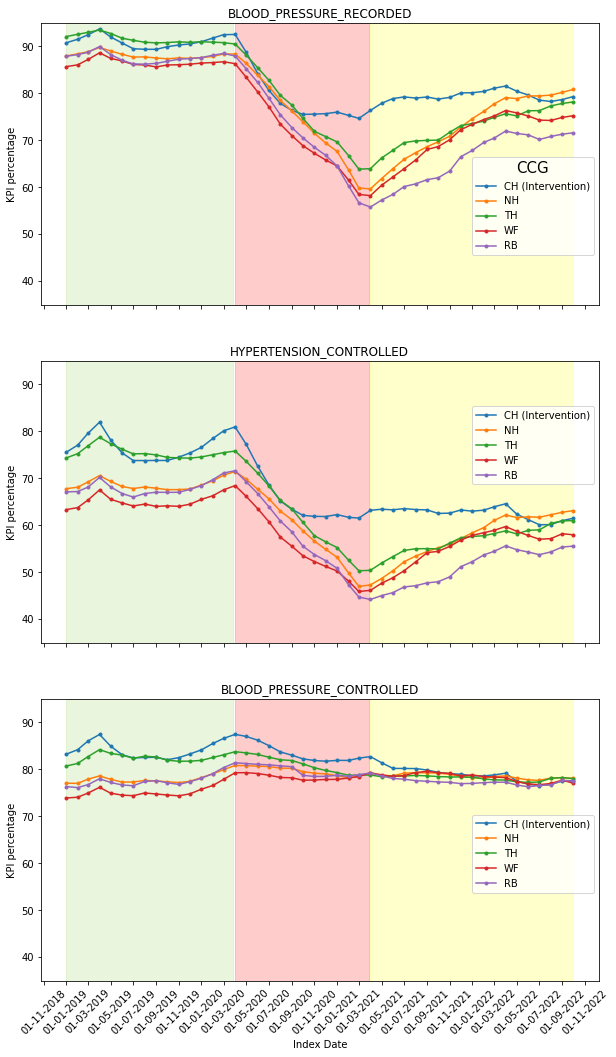

**Figure S2:** Indicator trends by study locality (CCG). In May 2020, the City and Hackney (CH) CCG launched a blood pressure recording initiative. Other localities are: NH: Newham, TH: Tower Hamlets, WF: Waltham Forest, RB: Redbridge.
