## Supplementary material for "COVID pandemic impact on hypertension management in North-East London: an observational cohort study using electronic health records": Figure S3

A. **BLOOD_PRESSURE_RECORDED** (blood pressure recorded within 12 months of the index date)

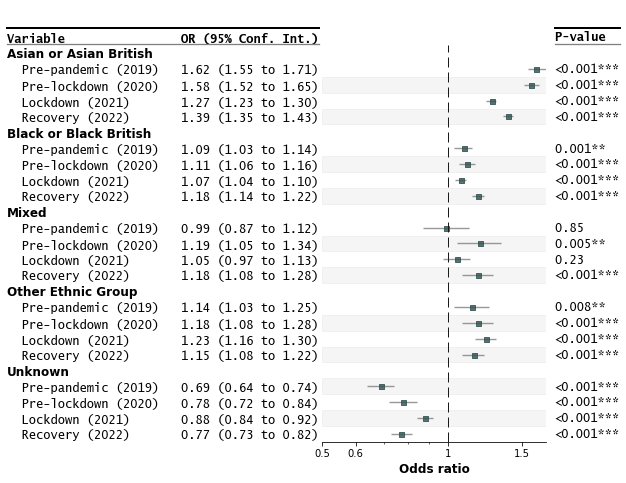

B. **HYPERTENSION_CONTROLLED** (most recent blood pressure on age-adjusted target; missing blood pressures are deemed uncontrolled)

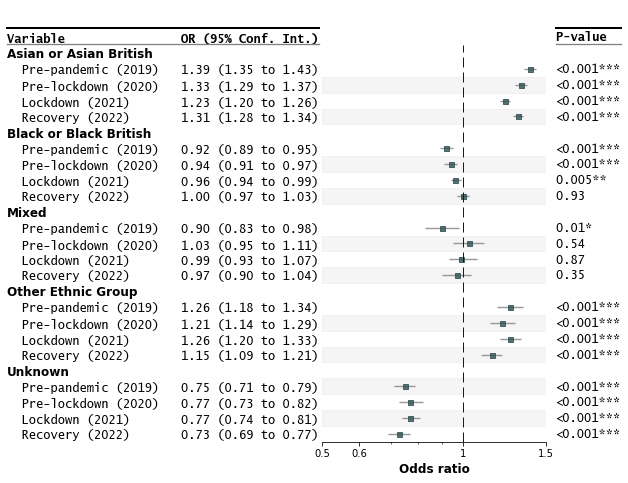

C. **BLOOD_PRESSURE_CONTROLLED** (most recent blood pressure on age-adjusted target; only individuals with a blood pressure recorded within last 12 months are considered)

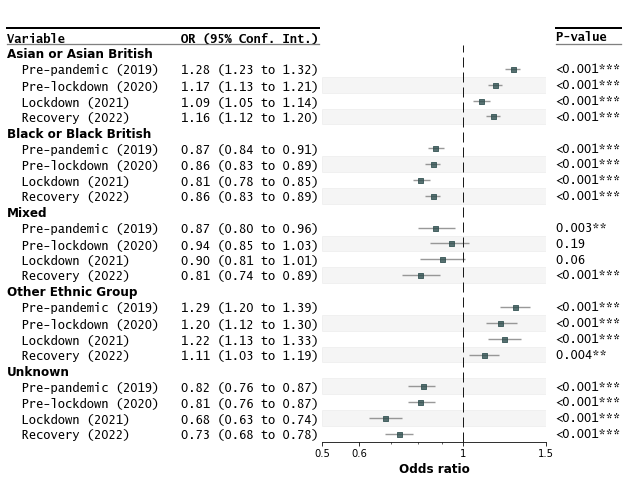

**Figure S3:** Impact of the pandemic on indicators of management of hypertension. Forest plots are shown for: A. BLOOD_PRESSURE_RECORDED, B. HYPERTENSION_CONTROLLED and C. BLOOD_PRESSURE_CONTROLLED. In each ethnicity group, the reference group is “White” in the same year (e.g. the Black/Black British ethnicity group in 2020 is 14% less likely to have controlled blood pressure than the White ethnicity group in 2020). The ORs are derived from a multivariate model adjusted for age, sex, IMD quintile and treatment intensity (see Supplementary Figures 4-6).
