## Supplementary material for "COVID pandemic impact on hypertension management in North-East London: an observational cohort study using electronic health records": Figure S4

A. **BLOOD_PRESSURE_RECORDED** (blood pressure recorded within 12 months of the index date)

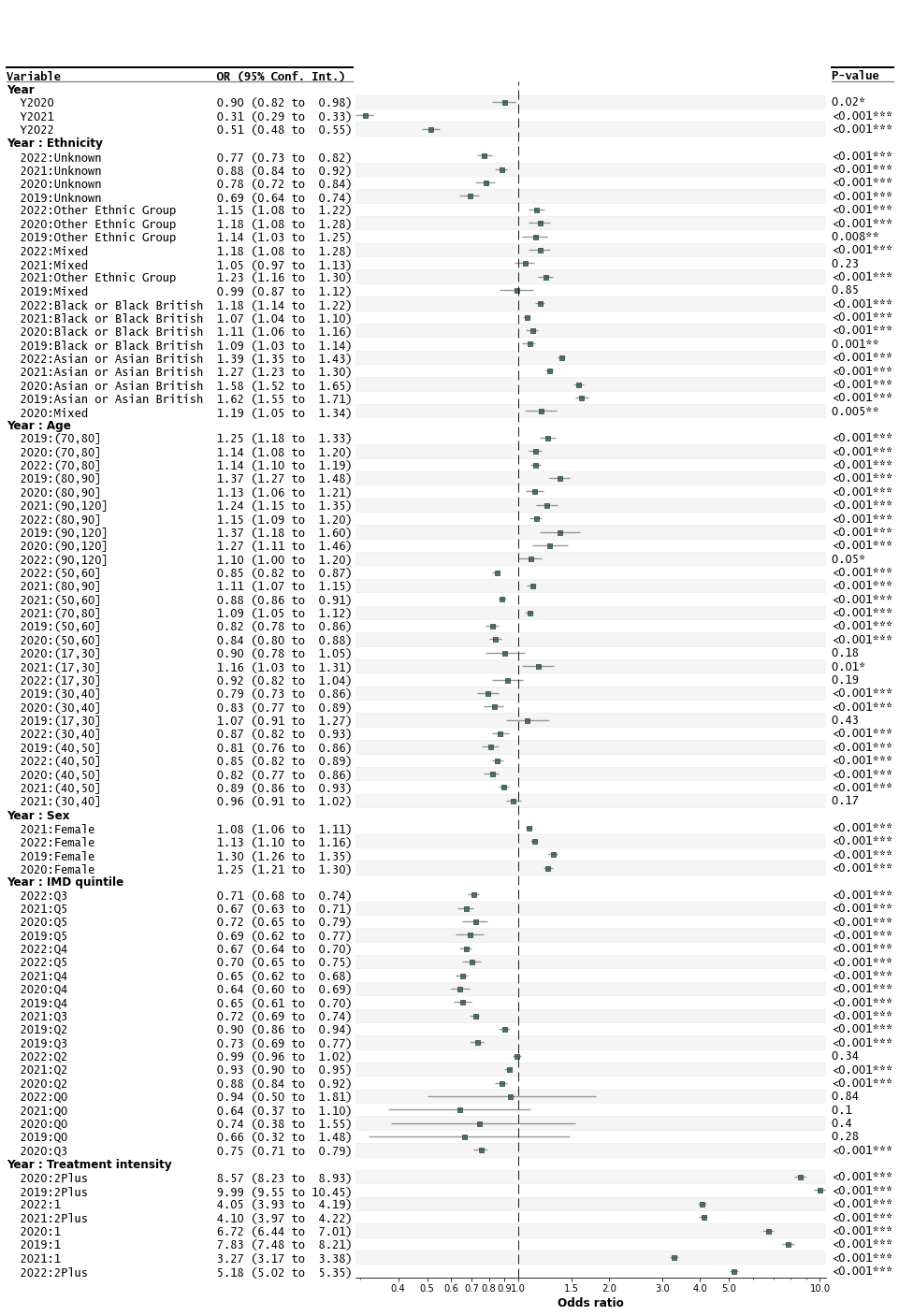

B. **HYPERTENSION_CONTROLLED** (most recent blood pressure on age-adjusted target; missing blood pressures are deemed uncontrolled)

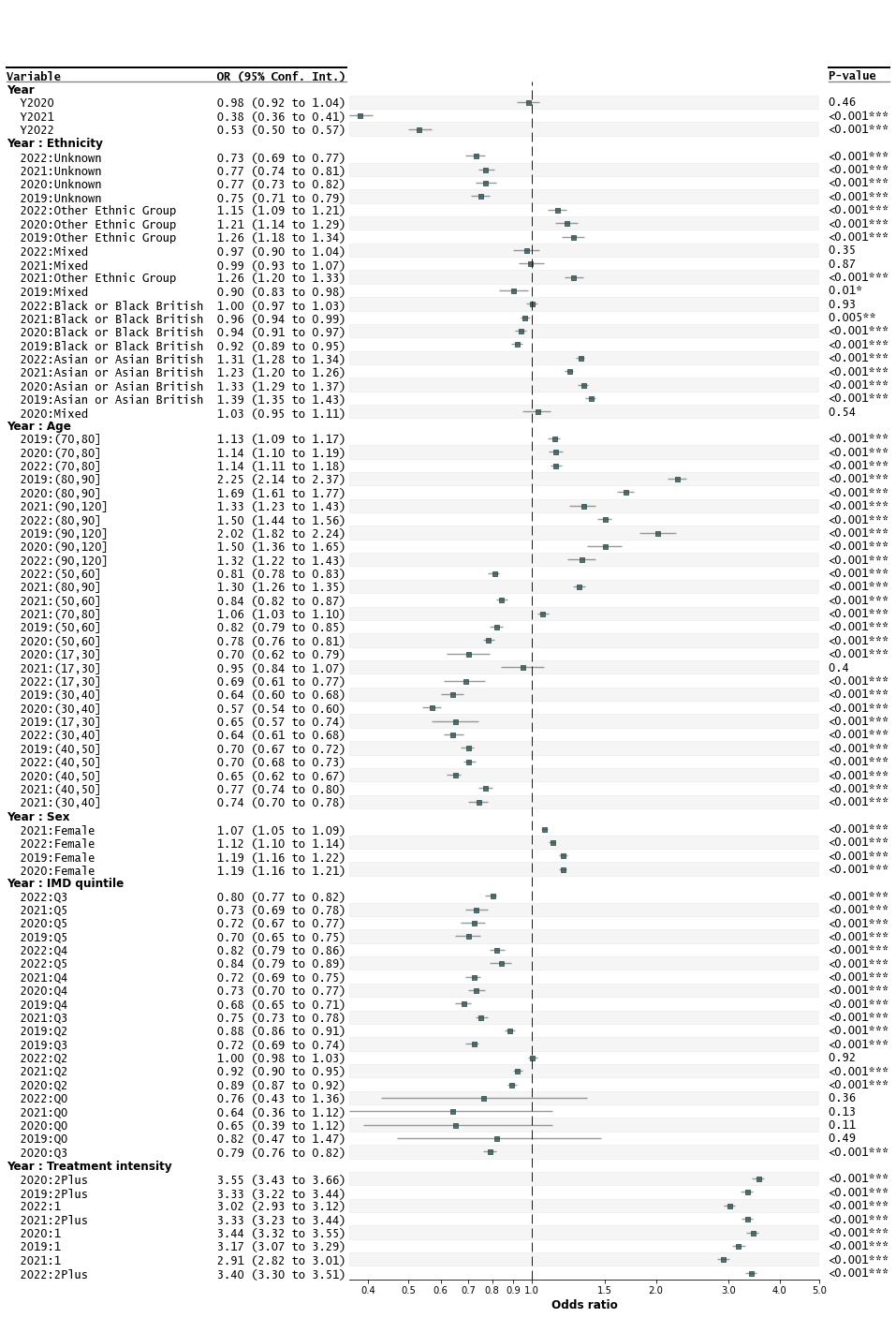

C. **BLOOD_PRESSURE_CONTROLLED** (most recent blood pressure on age-adjusted target; only individuals with a blood pressure recorded within last 12 months are considered)

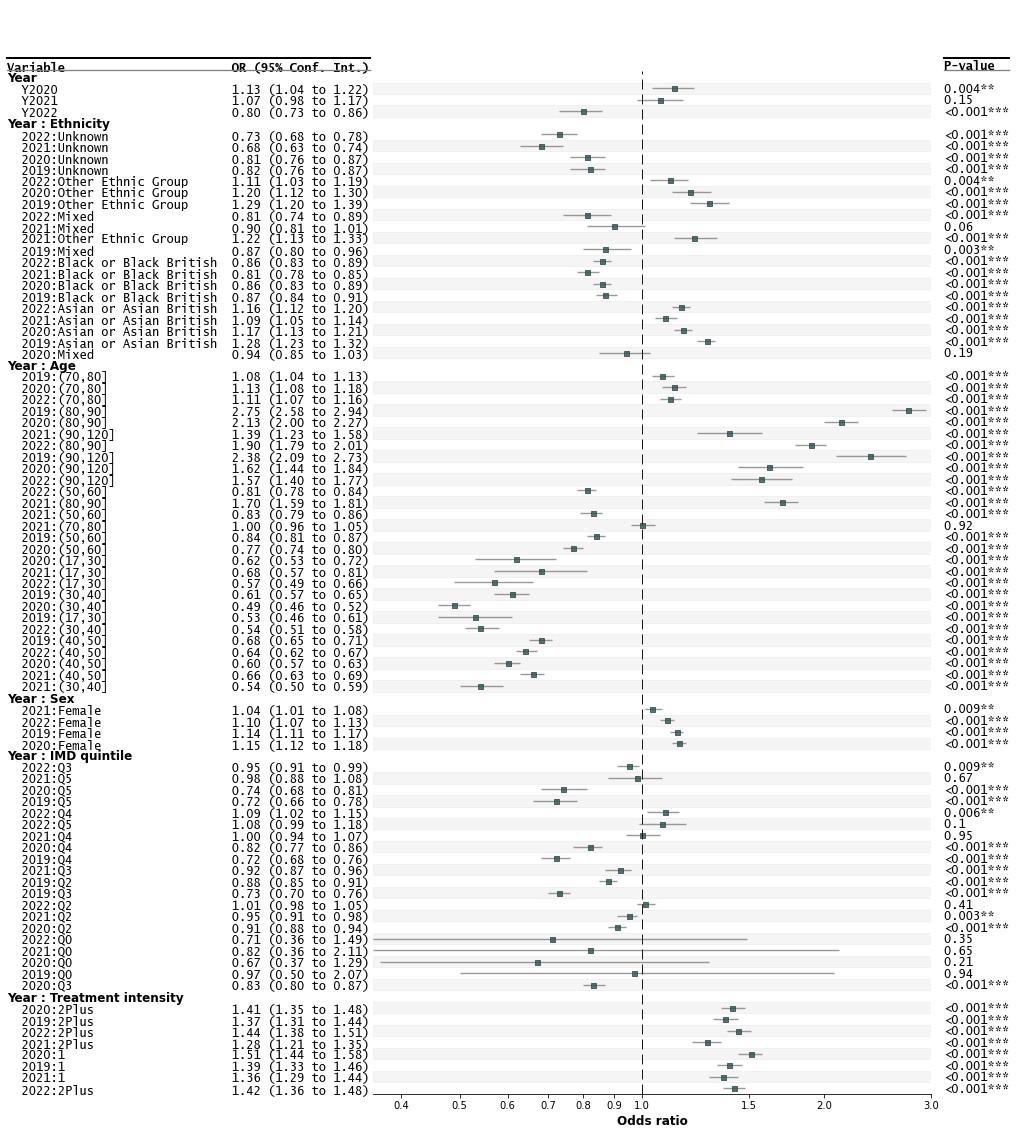

**Figure S4:** Full outcome of multivariable logistic regression analyses for the three study outcome indicators: A. BLOOD_PRESSURE_RECORDED, B. HYPERTENSION_CONTROLLED, C. BLOOD_PRESSURE_CONTROLLED. *: p-val <0.05; **: p-val<0.01; ***: p-val<0.001. *R model = OUTCOME ~ PANDEMIC_YEAR_GROUP + PANDEMIC_YEAR_GROUP:NHS_5_GROUP + PANDEMIC_YEAR_GROUP:AGE_GROUP + PANDEMIC_YEAR_GROUP:SEX_GROUP + PANDEMIC_YEAR_GROUP:QUINTILE_GROUP + PANDEMIC_YEAR_GROUP:NUMMED_GROUP*
