## Supplementary material for "COVID pandemic impact on hypertension management in North-East London: an observational cohort study using electronic health records": Table S1

| **Hypertension Code** | **Coding System** | **Description** | **Associated Read Code(s)** |
| --- | --- | --- | --- |
| G2 | Read | Hypertensive disease |  |
| G2_ | Read | Hypertensive disease |  |
| G20 | Read | Essential hypertension |  |
| G20.. | Read | Essential hypertension |  |
| G24 | Read | Secondary hypertension |  |
| G24.. (excl. G24z1) | Read | Secondary hypertension |  |
| G25 | Read | Stage 1 hypertension (NICE - Nat Ins for Hth Clin Excl 2011) |  |
| G25.. | Read | Stage 1 hypertension (NICE - Nat Ins for Hth Clin Excl 2011) |  |
| G26 | Read | Severe hypertension (Nat Inst for Health Clinical Ex 2011) |  |
| G28 | Read | Stage 2 hypertension (NICE - Nat Ins for Hth Clin Excl 2011) |  |
| G2y | Read | Other specified hypertensive disease |  |
| G2y.. | Read | Other specified hypertensive disease |  |
| G2z | Read | Hypertensive disease NOS |  |
| Gyu2 | Read | [X]Hypertensive diseases |  |
| Gyu2.. | Read | [X]Hypertensive diseases |  |
| 10441000006116 | SNOMED | Other specified hypertensive disease |  |
| 1201005 | SNOMED | Benign essential hypertension | G20.. |
| 1215744012 | SNOMED | Hypertensive disorder |  |
| 121910014 | SNOMED | Benign secondary renovascular hypertension |  |
| 131046010 | SNOMED | Malignant essential hypertension |  |
| 147988014 | SNOMED | Malignant secondary hypertension |  |
| 151171000006110 | SNOMED | Secondary malignant hypertension NOS |  |
| 1806071000006102 | SNOMED | Stage 1 hypertension | G2y.. |
| 1806071000006118 | SNOMED | Stage 1 hypertension |  |
| 1806081000006104 | SNOMED | Stage 2 hypertension | G2y.. |
| 1806081000006115 | SNOMED | Stage 2 hypertension |  |
| 1806141000006109 | SNOMED | Severe hypertension | G2y.. |
| 1806141000006113 | SNOMED | Severe hypertension |  |
| 194783001 | SNOMED | Secondary malignant renovascular hypertension | G24.. |
| 194785008 | SNOMED | Secondary benign hypertension | G24.. |
| 194788005 | SNOMED | Hypertension secondary to endocrine disorders | G24.. |
| 196353013 | SNOMED | Elevated blood pressure |  |
| 196354019 | SNOMED | Finding of increased blood pressure |  |
| 2164904016 | SNOMED | HTN - Hypertension |  |
| 2189411000000111 | SNOMED | Stage 1 hypertension (NICE - National Institute for Health and Clinical Excellence 2011) |  |
| 2189451000000110 | SNOMED | Severe hypertension (NICE - National Institute for Health and Clinical Excellence 2011) |  |
| 2193021000000110 | SNOMED | Severe hypertension |  |
| 2193031000000112 | SNOMED | Stage 1 hypertension |  |
| 2194941000000119 | SNOMED | Stage 2 hypertension (NICE - National Institute for Health and Clinical Excellence 2011) |  |
| 2211211000000110 | SNOMED | Stage 2 hypertension |  |
| 2335761000000116 | SNOMED | Stage 1 hypertension (NICE 2011) without evidence of end organ damage |  |
| 2335801000000114 | SNOMED | Stage 1 hypertension (NICE 2011) with evidence of end organ damage |  |
| 24184005 | SNOMED | Finding of increased blood pressure | G20.. |
| 2470030014 | SNOMED | Malignant secondary renovascular hypertension |  |
| 2470031013 | SNOMED | Benign secondary hypertension |  |
| 2478822013 | SNOMED | Secondary benign renovascular hypertension |  |
| 2532161014 | SNOMED | Hypertension secondary to endocrine disorder |  |
| 2671386015 | SNOMED | Hypertensive disorder; systemic arterial |  |
| 2920698012 | SNOMED | Blood pressure elevation |  |
| 299676019 | SNOMED | Secondary malignant renovascular hypertension |  |
| 299678018 | SNOMED | Secondary benign hypertension |  |
| 299681011 | SNOMED | Hypertension secondary to endocrine disorders |  |
| 3135013 | SNOMED | Benign essential hypertension |  |
| 31992008 | SNOMED | Secondary hypertension | G24.., Gyu2.. |
| 3763241013 | SNOMED | Endocrine hypertension |  |
| 38341003 | SNOMED | Hypertensive disease | G2_, G20.., G2y.., G2z, Gyu2.. |
| 389331000006110 | SNOMED | [X]Hypertension secondary to other renal disorders |  |
| 389341000006117 | SNOMED | [X]Hypertensive diseases |  |
| 413461000006118 | SNOMED | [X]Other secondary hypertension |  |
| 48146000 | SNOMED | Diastolic hypertension | G20.. |
| 490277011 | SNOMED | BP - High blood pressure |  |
| 490278018 | SNOMED | Systemic arterial hypertension |  |
| 490280012 | SNOMED | HBP - High blood pressure |  |
| 490281011 | SNOMED | HT - Hypertension |  |
| 490282016 | SNOMED | High blood pressure disorder |  |
| 490283014 | SNOMED | BP+ - Hypertension |  |
| 503982017 | SNOMED | Accelerated essential hypertension |  |
| 508416019 | SNOMED | Accelerated secondary hypertension |  |
| 523801000006119 | SNOMED | BP - hypertensive disease |  |
| 53452019 | SNOMED | Secondary hypertension |  |
| 56218007 | SNOMED | Systolic hypertension | G20.. |
| 59621000 | SNOMED | Essential hypertension | G20.. |
| 64168014 | SNOMED | Hypertensive disease |  |
| 64172013 | SNOMED | High blood pressure |  |
| 64173015 | SNOMED | Hypertensive vascular disease |  |
| 64174014 | SNOMED | Hypertensive vascular degeneration |  |
| 64176011 | SNOMED | Hypertension |  |
| 648911000006113 | SNOMED | Essential hypertension NOS |  |
| 73410007 | SNOMED | Secondary benign renovascular hypertension | G24.. |
| 78975002 | SNOMED | Malignant essential hypertension | G20.. |
| 80224019 | SNOMED | Diastolic hypertension |  |
| 843821000000102 | SNOMED | Stage 1 hypertension | G25.. |
| 843841000000109 | SNOMED | Severe hypertension | G26 |
| 846371000000103 | SNOMED | Stage 2 hypertension | G28 |
| 884121000006111 | SNOMED | Malignant hypertension |  |
| 89242004 | SNOMED | Malignant secondary hypertension | G24.. |
| 908631000000108 | SNOMED | Stage 1 hypertension (NICE 2011) without evidence of end organ damage | G25.. |
| 908651000000101 | SNOMED | Stage 1 hypertension (NICE 2011) with evidence of end organ damage | G25.. |
| 93494011 | SNOMED | Systolic hypertension |  |
| 99042012 | SNOMED | Essential hypertension |  |
| 99044013 | SNOMED | Idiopathic hypertension |  |
| 99046010 | SNOMED | Systemic primary arterial hypertension |  |
| 99047018 | SNOMED | Primary hypertension |  |
| **Hypertension resolved code** | **Coding System** | **Description** | **Associated Read Code(s)** |
| 21261 | Read | Hypertension resolved |  |
| 212K | Read | Hypertension resolved |  |
| 162659009 | SNOMED | Hypertension resolved | 21261, 212K |

Table S1: Hypertension and Hypertension resolved codes. Available from https://clinicalcodes.rss.mhs.man.ac.uk/medcodes/article/203/
