## Supplementary material for "COVID pandemic impact on hypertension management in North-East London: an observational cohort study using electronic health records": Table S2

| **Datum** | **Category** |
| --- | --- |
| Age | Patient Details |
| Gender | Patient Details |
| Lower Layer Area (2011) | Patient Details |
| Organisation Code | Patient Details |
| Ethnicity | Clinical Code |
| Hypertension | Clinical Code |
| Systolic BP | Clinical Code |
| Systolic BP | Date |
| Systolic BP | Value |
| Diastolic BP | Clinical Code |
| Diastolic BP | Date |
| Diastolic BP | Value |
| ACEi/ARB | Name, Dosage and Quantity |
| ACEi/ARB | Date of Issue |
| Beta Blocker | Name, Dosage and Quantity |
| Beta Blocker | Date of Issue |
| K-Sparing | Name, Dosage and Quantity |
| K-Sparing | Date of Issue |
| CCBs | Name, Dosage and Quantity |
| CCBs | Date of Issue |
| Thiazide Diuretic | Name, Dosage and Quantity |
| Thiazide Diuretic | Date of Issue |
| Centrally Acting HTs | Name, Dosage and Quantity |
| Centrally Acting HTs | Date of Issue |
| Alpha Blockers | Name, Dosage and Quantity |
| Alpha Blockers | Date of Issue |
| Loop Diuretic | Name, Dosage and Quantity |
| Loop Diuretic | Date of Issue |

**Table S2:** Patient data collected.
