## Supplementary material for "COVID pandemic impact on hypertension management in North-East London: an observational cohort study using electronic health records": Table S3

|  | **Total number of instances** | **Number of unique cohort individuals** |
| --- | --- | --- |
| ***Pre-processing dataset*** |  |  |
| Patient months | 7,514,995 | 224,329 |
| *Patient blood pressure recordings* | *7,286,480* |  |
| ***Patient instances excluded*** |  |  |
| Patients with sex recorded as “Unspecified” or “Unrecorded” | 38 | [SDL]* |
| ***Invalid blood pressure recordings (blood pressure reading excluded but instance not excluded)*** |  |  |
| Blood pressure recorded over a year prior to index date | 1,406,398 | 145,526 |
| Incomplete blood pressures (systolic blood pressure but no diastolic blood pressure and vice-versa) | 188 | [SDL] |
| Separately recorded blood pressure elements (i.e. date of systolic blood pressure different from date of diastolic blood pressure) | 2,240 | 391 |
| Diastolic blood pressure greater than or equal to systolic blood pressure | 343 | 104 |
| Unfeasible blood pressure (SBP < 70mmHg,  SBP ≥ 270mmHg, DBP < 40mmHg or DBP ≥ 150mmHg) | 2,348 | 409 |
| ***Post-processing dataset*** |  |  |
| Patient months | 7,514,957 | 224,329* |
| *Patient months excluded* | *38* |  |
| Patient BPs | 5,874,925 | 215,219 |
| *Patient BPs excluded* | *1,411,555* |  |

**Table S3:** Cohort processing. BP = Blood pressure; SBP = Systolic Blood Pressure; DBP = Diastolic Blood Pressure; SDL = statistical disclosure limitation (number in group not disclosed for groups with fewer than 100 individuals). *the overall number of individuals is not affected as these individuals only have Sex recorded as unspecified/unrecorded for a part of the study
