## Supplementary material for "COVID pandemic impact on hypertension management in North-East London: an observational cohort study using electronic health records": Table S4

|  | **2019** | | **2020** | | **2021** | | **2022** | |
| --- | --- | --- | --- | --- | --- | --- | --- | --- |
|  | OR | 95% CI | OR | 95% CI | OR | 95% CI | OR | 95% CI |
| **BLOOD_PRESSURE_RECORDED** | | | | | | | | |
| 1 antihypertensive | 7.83 | 7.48-8.21 | 6.72 | 6.44-7.01 | 3.27 | 3.17-3.38 | 4.05 | 3.93-4.19 |
| 2+ antihypertensives | 9.99 | 9.55-10.45 | 8.57 | 8.23-8.93 | 4.10 | 3.97-4.22 | 5.18 | 5.02-5.35 |
| **HYPERTENSION_CONTROLLED** | | | | | | | | |
| 1 antihypertensive | 3.17 | 3.07-3.29 | 3.44 | 3.32-3.55 | 2.91 | 2.82-3.01 | 3.02 | 2.93-3.12 |
| 2+ antihypertensives | 3.33 | 3.22-3.44 | 3.55 | 3.43-3.66 | 3.33 | 3.23-3.44 | 3.40 | 3.30-3.51 |
| **BLOOD_PRESSURE_CONTROLLED** | | | | | | | | |
| 1 antihypertensive | 1.39 | 1.33-1.46 | 1.51 | 1.44-1.58 | 1.36 | 1.29-1.44 | 1.44 | 1.38-1.51 |
| 2+ antihypertensives | 1.37 | 1.31-1.44 | 1.41 | 1.35-1.48 | 1.28 | 1.21-1.35 | 1.42 | 1.36-1.48 |

**Table S4:** Effect of treatment intensity on study indicators. The reference group (OR = 1.00) are untreated patients (i.e. patient on no antihypertensive medication). **All p-values are <0.001.**
